# Glycomic Analysis Identifies Pre-Vaccination Markers of Response to Influenza Vaccine, Implicating the Complement Pathway

**DOI:** 10.1101/2022.02.09.22270754

**Authors:** Rui Qin, Guanmin Meng, Smruti Pushalkar, Michael A. Carlock, Ted M. Ross, Christine Vogel, Lara K. Mahal

## Abstract

Response to vaccination can vary significantly from person to person. A key to improving vaccine design and vaccination strategy is to understand the mechanism behind this variation. The role of glycosylation, a critical modulator of immunity, is unknown in determining vaccine responses. To gain insight into the association between glycosylation and vaccine-induced antibody levels we profiled the pre- and post-vaccination serum protein glycomes of 160 Caucasian adults receiving the FLUZONE™ influenza vaccine during the 2019-2020 influenza season. Using lectin microarrays, we observed that pre-vaccination levels of Lewis A antigen (Le^a^) are significantly higher in people who did not mount significant antibody responses, when compared to responders. Glycoproteomic analysis showed that Le^a^-bearing proteins are enriched in complement activation pathways, suggesting a potential role of glycosylation in tuning the activities of complement proteins, which may be implicated in mounting vaccine responses. We also observed post-vaccination increases in sialyl Lewis X antigen (sLe^x^) and decreases in high mannose glycans among high responders, which were not observed in non-responders. This data suggests that the immune system may actively modulate glycosylation as part of its effort to establish effective protection post-vaccination.

## INTRODUCTION

Vaccination is a powerful means to prevent infection and the development of severe disease. However, the response to vaccination varies across populations in ways that we have yet to understand.^1^ Vaccines against influenza, a major respiratory illness causing millions of hospitalizations and hundreds of thousand deaths worldwide each year, are perhaps the most widely studied, in part because vaccinations are administered annually.^2–4^ In the past decade, research has identified several potential biomarkers associated with influenza vaccine response. These include hormones, cytokines, pre-existing antibodies and pre-existing T cell populations.^5–7^

Glycosylation is an underexplored modulator of immune response to influenza vaccines, in spite of the multifaceted roles glycans have been found to play in immunity.^8,9^ For example, antibody glycosylation has been found to alter the ability of antibodies to activate the immune system and changes their circulation time in sera.^10–12^ Two pioneering studies conducted in influenza-vaccinated cohorts revealed changes in *N*-glycan sialylation, galactosylation and fucosylation on influenza-specific IgG post-vaccination.^13,14^ However, the level of viral binding antibodies produced (antibody response) in association with the glycosylation of these antibodies was not examined. Sera contain a host of other proteins involved in immunity beyond IgG, including innate immune lectins, complement factors, and other glycoproteins. Comprehensive profiling of the serum glycome may provide valuable insights into the mechanisms underlying vaccine response and identify new glycan-based markers.

Herein we analyze pre- and post-vaccination serum glycomes and associated antibody responses in a cohort of 160 Caucasian adults vaccinated with the widely used FLUZONE™ influenza vaccine. Glycomic analysis was performed using our dual-color lectin microarray technology which has been used in a wide variety of studies including analysis of exosomes, host-response to influenza, and identification of cancer drivers in human tissues.^15–21^ We found that individuals who lacked response to vaccination had high levels of the blood group epitope Lewis A (Le^a^). Glycoproteomic analysis identified multiple complement components and other immune-related proteins marked with this epitope, which was enriched in non-responders. We also observed post-vaccination upregulation of sialyl Lewis X (sLe^x^) and down-regulation of high mannose glycans in high responders, neither of which was observed in non-responders. Our data suggest that glycosylation may help predict immune response to vaccine and that specific glycoforms of immune proteins may be important in tuning vaccination response.

## METHODS

### Cohort Recruitment

160 Caucasian adults were enrolled at the University of Georgia Clinical and Translational Research Unit (Athens, Georgia, USA) from September 2019 to February 2020. All volunteers were enrolled with written, informed consent. Participants were excluded if they already received the seasonal influenza vaccine. Other exclusion criteria included acute or chronic conditions that would put the participant at risk for an adverse reaction to the blood draw or the flu vaccine (*e*.*g*., Guillain-Barré syndrome or allergies to egg products), or conditions that could skew the analysis (*e*.*g*., recent flu symptoms or steroid injections/medications). All participants received a FLUZONE™ (Sanofi Pasteur, Lyon, France), seasonal inactivated influenza vaccine. Most received a quadrivalent, standard dose formulation made up of 15μg HA per strain of A/H1N1 (A/Brisbane/02/2018), A/H3N2 (A/Kansas/14/2017), B/Yamagata (B/Phuket/3073/2013) and B/Victoria (B/Colorado/6/2017-like strain).

### Hemagglutination Inhibition Assays

Blood samples were collected from participants prior to vaccination (d0) and again post-vaccination at d28 (22-35 days). Hemagglutinin inhibition (HAI) assays were performed with serum from each participant at d0 and d28. Sera was used at a starting concentration of 1:10 following treatment with a receptor-destroying enzyme (RDE) (Denka Seiken, Tokyo, Japan) to inactivate non-specific inhibitors. RDE-treated sera (25 μL), including positive and negative controls, were serially diluted in PBS (2.67 mM KCl, 1.47 mM KH_2_PO_4_, 8.10 mM Na_2_HPO_4_, 138 mM NaCl, pH = 7.4, same hereinafter) two-fold across 96-well V-bottom microtiter plates. An equal volume of influenza virus (25 μL), which was adjusted beforehand via hemagglutination (HA) assay to a concentration of 8 hemagglutination units (HAU) per 50 μL, was added to each well and incubated at room temperature for 20 minutes. Finally, 0.8% turkey red blood cells (Lampire Biologicals, Pipersville, Pennsylvania, USA) in PBS were added, plates mixed by agitation, and then incubated at room temperature for 30 minutes. The HAI titer was determined by the reciprocal dilution of the last well that contained non-agglutinated RBCs.

### Definition of Antibody Responses

Response scores for each strain of influenza were calculated based on the fold changes of antibody titers (d28 titer / d0 titer). For each strain, antibody response was scored in the following steps: i) calculate the initial score by taking the logarithmic (base 2) value of the titer fold change; ii) change the score to zero if the d28 antibody titer is lower or equal to 20, a conventional cut-off for effective protection;^22,23^ iii) change the score to 4 if the initial score is greater than 4 (*i*.*e*., an over 16-fold increase in titer). This is to prevent the total response score (see below) from being biased towards one single strain. This strain-specific score was used to categorize the participants into three response groups: high responders (score ≥ 2), low responders (1 ≤ score < 2) and non-responders (score < 1). The total response score is the sum of the four strain-specific scores. Similarly, total response scores were used to define overall high responders (score ≥ 8), overall low responders (4 ≤ score < 8) and overall non-responders (score < 4).

### Fluorescent Labelling of Serum Proteins

Total protein concentrations of serum samples were measured with *DC*™ protein assay kit (Bio-Rad Laboratories, Hercules, California, USA). Each volunteer serum sample was fluorescently labelled with Alexa Fluor 555™ NHS ester (Thermo Fisher Scientific, Waltham, Massachusetts, USA). First, 10 μg of total protein was diluted in PBS to 27 μL. The pH of the solution was adjusted with 3 μL of 1M sodium bicarbonate. Then 0.21 μL of a stock solution (10 mg/mL) of Alexa Fluor™ 555 NHS ester was added to the mixture. The reaction lasted for 1 hour in the dark at room temperature. Unconjugated dye molecules were then removed by Zeba™ Dye and Biotin Removal Filter Plates (Thermo Fisher Scientific, Waltham, Massachusetts, USA). The reference material, NIST human serum 909c (Millipore Sigma, Darmstadt, Germany), was fluorescently labelled with Alexa Fluor 647™ NHS ester (Thermo Fisher Scientific, Waltham, Massachusetts, USA) similarly. The amounts of reagents were scaled linearly to the starting protein amount (4 mg). Finally, each Alexa Fluor 555-labelled sample (10 μg of total protein) was mixed with a proper volume of Alexa Fluor™ 647-labelled reference material containing the same amount of protein, and the final volume was adjusted to 50 μL with PBS.

### Dual-color Lectin Microarray

Lectin microarray slides were fabricated as previously described.^24^ A list of probes printed on the array is in Supporting Information, Table S1. The print was quality controlled as previously described.^24^ Prior to hybridization, each dual-color mixture was diluted with 50 μL 0.2% PBST (PBS with 0.2% Tween-20, v/v). Each mixture was then allowed to hybridize with the arrays for 1 hour in the dark at room temperature. Arrays were washed twice with 0.005% PBST for 5 minutes and once with PBS for 5 minutes. The slides were briefly rinsed with ultrapure water and dried. Florescence signals were obtained with Genepix™ 4400A fluorescence slide scanner (Molecular Devices, San Jose, California, USA) in the 532 nm channel and the 635 nm channel that correspond to the excitation/emission profiles of Alexa Fluor™ 555 and Alexa Fluor™ 647, respectively. Raw fluorescence signal and background signal of each spot were generated by the Genepix Pro™ 7 software (Molecular Devices, San Jose, California, USA), which were further processed and analyzed with a custom script as previously described.^16^ Heatmaps and volcano plots were generated with R. Lectin microarray data is available at Synapse.org (doi: 10.7303/syn26956958).

### Lectin/Antibody Affinity Pulldown

BambL (80 μg, expressed in-house) or anti-Le^a^ (80 μg, Abcam, Cambridge, United Kingdom) was immobilized on columns using AminoLink™ Plus Micro Immobilization Kit (Thermo Fisher Scientific, Waltham, Massachusetts, USA) as per manufacturer’s protocol. Coupling was carried out at 4°Covernight with gentle agitation. For the beads-only controls, PBS was added to the columns instead of BambL/anti-Le^a^ in the coupling step.

For protein identification by proteomics, a serum pool was prepared by combining 10 μL of each day 0 serum sample. The pooled serum was incubated at 54°Cfor 1 hour to inactivate proteases prior to the pulldown experiments. For BambL pulldown, 10 μL of pooled serum was diluted in PBS to 200 μL and incubated on the column for 1 hour at room temperature with gentle agitation. The column was washed with 300 μL PBS three times (5 minute per wash with gentle agitation). The column was eluted with 200 μL 50mM methyl α-L-fucopyranoside (TCI America, Portland, Oregon, USA) in PBS. For anti-Le^a^ pulldown, 50 μL of pooled serum was diluted in PBS to 400 μL and incubated on the column for 1 hour at room temperature with gentle agitation. The column was washed with 300 μL PBS three times (5 minute per wash with gentle agitation) before being eluted with 100 μL 0.1M glycine (pH = 2.8). The eluate was immediately neutralized with 30 μL 0.5M Tris (pH = 8.5). The protocol for preparing the six BambL-pulldown samples for western blotting is the same as the protocol for glycoproteomics except that the columns were incubated with 200 μg serum protein diluted in 100 μL PBS, washed with 100 μL PBS and eluted with 80 μL elution buffer.

### Peptide preparation and LC-MS/MS analysis

The enriched samples were incubated at 95°C for 10 min. 1 μg/μL of sequencing grade-modified trypsin (Promega, Madison, Wisconsin, USA) was added to samples and overnight at 37°C with gentle agitation. Digestion was quenched by pH <4.0 using 2.5% trifluoroacetic acid (TFA). Samples were subsequently desalted using Pierce C18 spin tips (Thermo Fisher Scientific, Waltham, Massachusetts, USA) as per manufacturer’s protocol. The peptides were eluted using aqueous buffer with 60% acetonitrile (ACN) and 0.1% formic acid (FA). The samples were dried, and peptides resuspended in 10 µl of buffer (0.1% FA in 5% ACN).

Each sample (∼3 μL) was loaded onto Acclaim PepMap 100 trap column (75 μm x 2 cm) nanoViper, attached to an EASY-spray analytical column (PepMap RSLC C18, 2 μm, 100Å, 75 μm ID x 50 cm) in an EASY nano-LC 1000 liquid chromatography instrument (Thermo Scientific). Chromatography solvent A consisted of LC-MS grade water with 0.1% FA, and solvent B of 80% acetonitrile with 0.1% FA. The 155 min gradient consisted of: 2-5% of solvent B for 5 min, 5-25% for 110 min, 25-40% for 25 min, 40-80% for 5 min, 80-95% for 5 min, followed by 95-5% for 5 min. Mass spectrometry data was collected in data dependent mode on an Orbitrap Eclipse mass spectrometer (Thermo Fisher Scientific, Waltham, Massachusetts, USA). The MS1 spectra were recorded with a resolution of 240,000, AGC target of 1e6, with maximum injection time of 50 ms, and a scan range of 400 to 1500 m/z. The MS2 spectra were collected using quadrupole isolation mode, AGC target of 2e4, maximum injection time of 18 ms, one microscan, 0.7m/z isolation window, collision energy of 27%, excluding ions of charge state <+2 and >+7.

Spectra were searched against the Uniprot human fasta sequence database (UP000005640, downloaded on July 24, 2020) using the MaxQuant software (version 1.5.5.1) with default settings, including 2 missed cleavages, first search with peptide tolerance of 20 ppm and for the main search with peptide tolerance of 4.5 ppm. Carbamidomethylation of Cysteine was set as a static modification. The false discovery rates for peptide and protein identifications were both set to 0.01. Oxidation of Met and acetylation of the protein N terminus were the allowed variable modifications, and proteins were quantified using the Label Free Quantification (LFQ) option.

A protein was identified as a positive binder if the enriched sample (E) and the corresponding control sample (C) satisfied the following: i) the sum of log_10_-transformed LFQ intensities of this protein in E and C was > 3; and ii) the difference of log_2_-transformed LFQ intensities of this protein between E and C was > 2 (E-C). The remaining proteins were searched in GlyGen^25^, a database that compiles the experimental evidence for glycosylation of proteins. Proteins without any experimental evidence for glycosylation or solely with experimental evidence for *O*-GlcNAcylation were removed, as they are not the targets of interest of the pulldown experiments.

### Pathway Enrichment Analysis

Gene Ontology (GO) enrichment analysis was performed with PANTHER Overrepresentation Test (Released 20210224).^26,27^ The input analyzed lists are the lists of glycoproteins identified in BambL or anti-Le^a^ pulldowns (Supporting Information, Table S2 and Table S3). A full list of plasma proteins was used as the reference list.^28^ “GO biological process complete”, “Fisher’s Exact” and “Calculate False Discovery Rate” were selected as the annotation data set, test type and correction method, respectively.

### Western Blotting

All steps were performed at room temperature. 20 μg of pulldown samples or input serum samples were resolved by 4-20% SDS/PAGE and transferred to a nitrocellulose membrane. Total protein was stained with Revert™ 700 Total Protein Stain Kit (LI-COR, Lincoln, Nebraska, USA) as per manufacturer’s instruction. After the total protein stain was erased, the membrane was blocked with blocking buffer [PBS with 3% (w/v) BSA and 0.05% (v/v) Tween-20] for 1 hour. Then the membrane was incubated with primary antibody working solution [rabbit anti-C4BPA (Abcam, Cambridge, United Kingdom), diluted to 0.5μg/ml in blocking buffer] for 1 hour, washed with PBST three times for 5 minutes per wash, incubated with secondary antibody working solution [goat anti-rabbit IgG CF™640R conjugate (Millipore Sigma, Darmstadt, Germany), diluted to 0.1μg/ml in blocking buffer] for 15 minutes and washed with PBST three times for 5 minutes per wash before imaging.

## RESULTS AND DISCUSSION

### Lectin Microarray Analysis Shows Clear Glycomic Differences Between High and Non-Responders Prior to Influenza Vaccination

To evaluate whether the glycome varies in individuals with differing vaccine response, we characterized the serum glycomes of 160 Caucasian adults who received FLUZONE™ quadrivalent vaccines in the 2019-2020 flu season in a medical facility in Georgia, USA. The FLUZONE™ vaccine is composed of four inactivated strains of influenza virus, including two type A strains (A/Brisbane/02/2018 (subtype H1N1), A/Kansas/14/2017 (subtype H3N2)) and two type B strains (B/Phuket/3073/2013 (Yamagata lineage) and B/Colorado/6/2017-like (Victoria lineage)). Pre-vaccination sera of participants were collected on the day of vaccination (d0). Post-vaccination sera were collected approximately four weeks post vaccination (d28). Antibody titers were determined via serum hemagglutination inhibition (HAI) assays, and the resulting log_2_ fold change (d28/d0) was used to calculate a response score for each strain (**Scheme 1** and **Methods**). To assess the overall antibody response across all four strains, we calculated a total response score by summing the individual response scores. Participants were categorized into high responders (total score ≥ 8, *N* = 67), low responders (4 ≤ total score < 8, *N* = 39), and non-responders (total score < 4, *N* = 54).

**Scheme 1.**
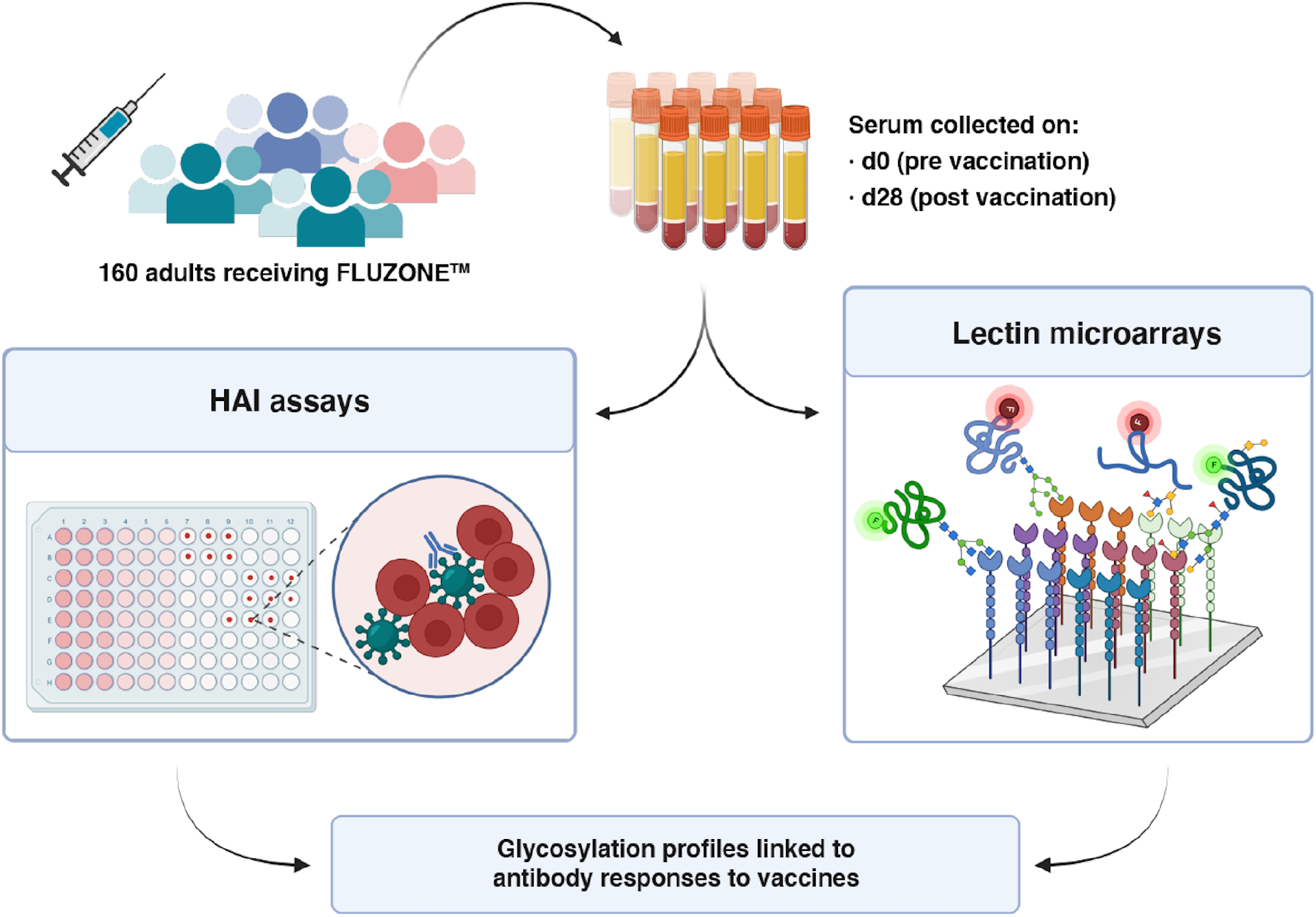
Workflow of integrated analysis. Hemagglutination inhibition assays (HAI) and lectin microarray assays were run on sera collected pre- (day 0, d0) and post- (day 28, d28) vaccination.

To analyze the glycome, we used our dual-color lectin microarray technology (**Scheme 1**).^15^ Lectin microarrays, which utilize well-characterized glycan-binding probes to identify glycomic changes at the substructure level, have been used to identify glycans driving cancer progression and metastasis,^19–21^ involved in exosome biogenesis,^16,29^ and associated with influenza severity.^17,18^ For this study, the probes included 68 lectins and 14 carbohydrate-binding antibodies. In addition, we printed protein A, protein G and protein L to assess serum immunoglobulin levels. In brief, each sample was labelled with Alexa Fluor 555. A serum standard (human serum 909c, NIST) was labelled with the orthogonal dye, Alexa Fluor 647, and used as the biological reference. Equal amounts (10 μg) of sample and reference were incubated on each array. Data was analyzed as previously described.^16^ An annotated heatmap of the pre-vaccination glycomic profiles with response score is shown in Figure 1.

**Figure 1.**
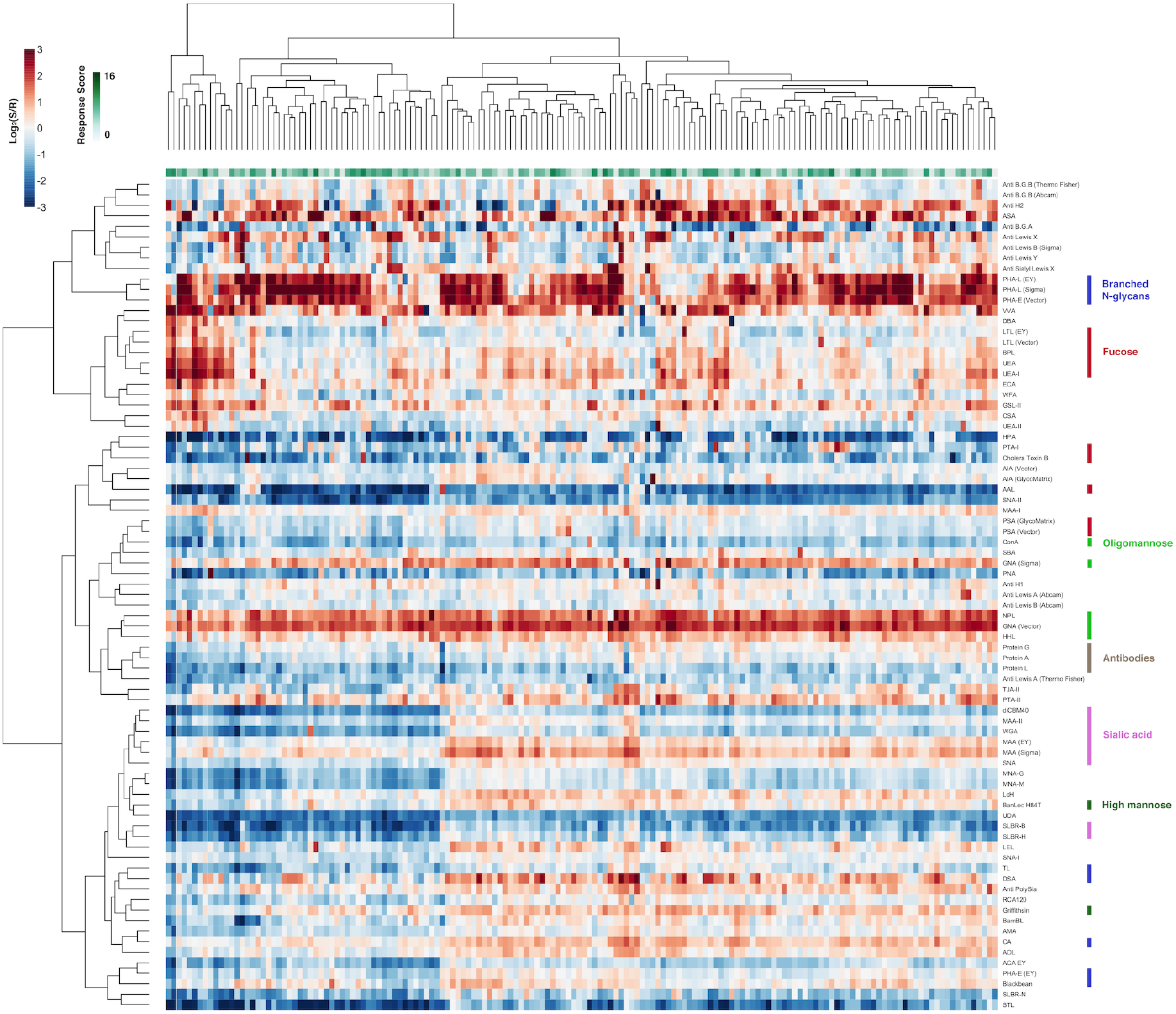
Heatmap of lectin microarray data for d0 serum samples. Columns represent the participants and rows represent the probes. Color of cells represent the normalized log_2_ ratios (Sample signal (S)/Reference signal (R)). Total response scores are annotated with a green-white sliding scale bar on the top of the heatmap.

To clearly identify glycan epitopes that might be predictive of vaccine response, we compared the glycomes at d0 of high responders to non-responders. We observed a clear pattern of glycan motifs associated with lack of response to the vaccine. In comparison to high responders, people with poor antibody response exhibited significantly higher binding to fucosylated Type I LacNAc antigens (**Figure 2**, probes: BambL, anti-Le^a^, anti-H1). The specificity of the anti-Le^a^ antibody overlaps with the *Burkholderia ambifaria* lectin (BambL), a pan-Lewis antigen binder that binds Le^a^, providing additional confirmation of the Le^a^ signature.^30^

**Figure 2.**
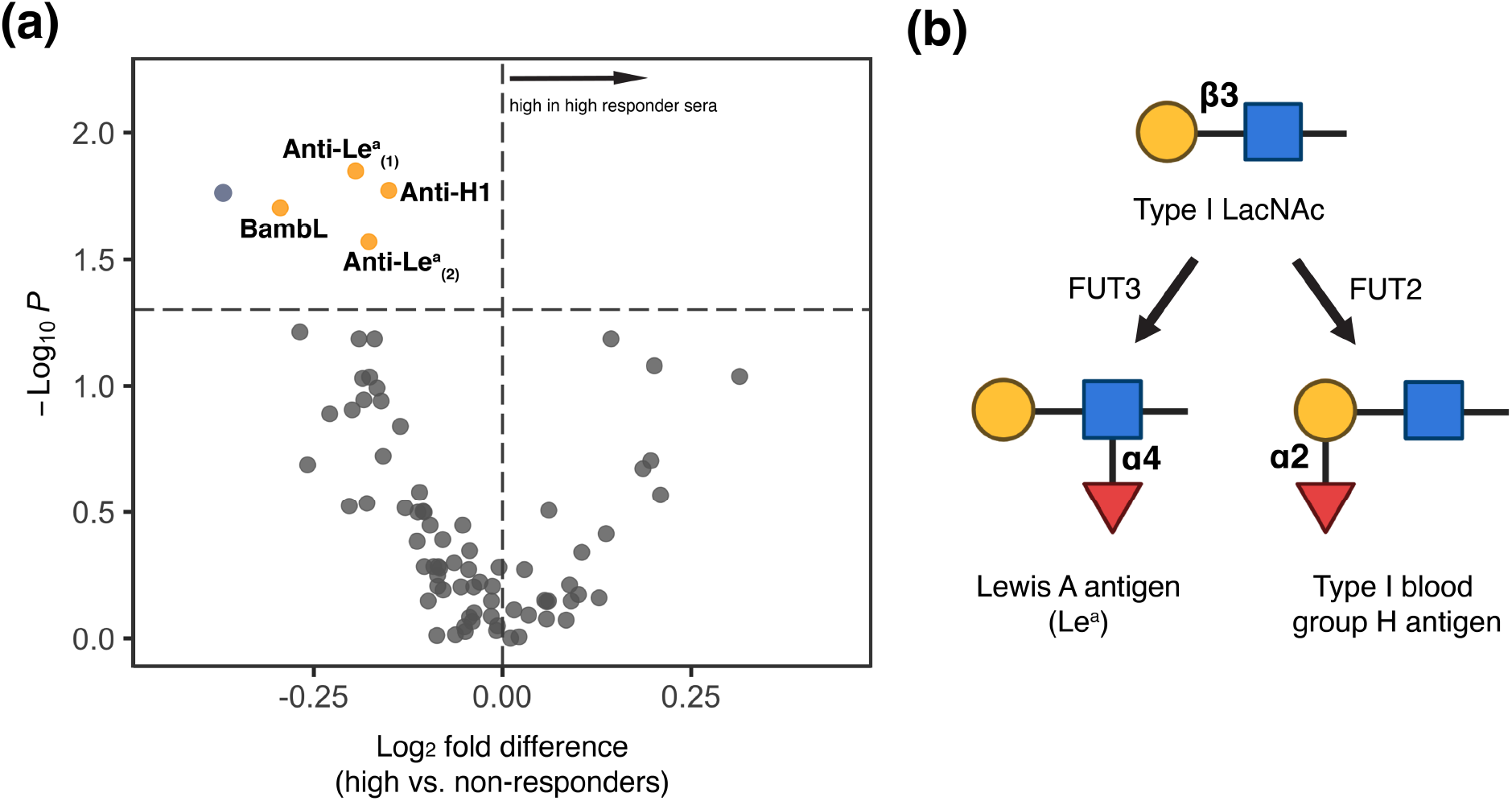
**(a)** Volcano plot comparing lectin microarray data for high responders (*N* = 65) and non-responders (*N* = 54) pre-vaccination. Mann–Whitney U test was used to determine *p-*values. Probes with *p* < 0.05 are colored in yellow. BambL: *Burkholderia ambifaria* lectin; Anti-Le^a^_(1)_ : anti-Lewis A, Invitrogen; Anti-Le^a^ _(2)_: anti-Lewis A, Abcam. Anti-H1: anti-type I blood group H (O), Invitrogen. **(b)** Partial biosynthetic routes of Lewis A antigen and type I blood group H (O) antigen. Glycans are shown in the Symbolic Nomenclature for Glycomics (SNFG). Symbols are defined as follows: galactose (yellow ●), N-acetylglucosamine (blue ■), fucose (red ▲). FUT2: Galactoside alpha-(1,2)- fucosyltransferase 2; FUT3: 3-galactosyl-N-acetylglucosaminide 4-alpha-L-fucosyltransferase.

Immune response to influenza has been found to be strain-dependent.^31,32^ The FLUZONE™ vaccine contains four different strains of influenza (two type A and two type B). To assess the impact of strain on the glycomic association with antibody response, we compared the high-responders and non-responders for each strain (high response: score ≥ 2, non-response: score < 1; **Supporting Information, Figure S1-S4**). We observed higher binding to BanLec^H84T^, a high mannose binding probe (Man_7_-Man_9_), for influenza B strains in participants categorized as non-responders **(Supporting Information, Figure S3 and S4)**. High mannose glycans in human sera have been associated with progression of breast cancer, hypercholesterolemia and nephropathy.^33–35^ In line with our previous analysis, we also observe higher binding to Le^a^ probes (anti-Le^a^, BambL) in non-responders for three of the four strains. Only the B/Victoria (B/Colorado/6/2017-like) strain did not show this association.

Le^a^ lacks well-characterized roles in immunity. Non-secretors (FUT2 deficient, **Figure 2b**) have higher levels of secreted Le^a^ in their blood (Le^a^ phenotype)^36^. Similar to our findings, a study among children vaccinated against norovirus showed association of this high Le^a^ phenotype with low seroconversion rates.^37^ Together with our data, this suggests an association between Le^a^ antigens and vaccine response and a heretofore unknown role for this glycan in immunity.

### Serum sLe^x^ is Upregulated and High-mannose is Downregulated in High Responders Post-vaccination

To assess whether changes in sera glycosylation as a function of immunity are observed post-vaccination, we generated glycomic profiles of the day 28 sera (**Figure 3a**). In general, the d28 glycomes significantly correlated with the d0 glycomes of the same individuals, indicating remarkable stability of the overall serum glycome post-vaccination compared to pre-vaccination (median correlation coefficient = 0.91, **Supporting Information, Figure S5**). To identify vaccine-induced changes in specific glycans, we next compared paired pre- (d0) and post- (d28) vaccination glycomes. We observe a loss of high mannose glycans post-vaccination (Griffithsin, BanLec^H84T^, HHL, **Supporting Information, Figure S6)**. This loss is clear in high responders but not in non-responders (**Figure 3b and Supporting Information, Figure S7 and Figure S8**). We also observe upregulation of Lewis X (Le^x^) antigens. This effect is seen in both high- and non-responders, with the non-responder group showing greater increase. In contrast, sialyl-Lewis X (sLe^x^) antigens are higher at d28 in high-responders, but this association is not observed in non-responders (**Figure 3c and Supporting Information, Figure S7 and Figure S8**). No other clear shifts in glycan levels were identified. Interestingly, total serum immunoglobulins seemed to decrease slightly in non-responders, as protein A, protein G and protein L all had lower binding to post-vaccination sera (**Supporting Information, Figure S8**).

**Figure 3.**
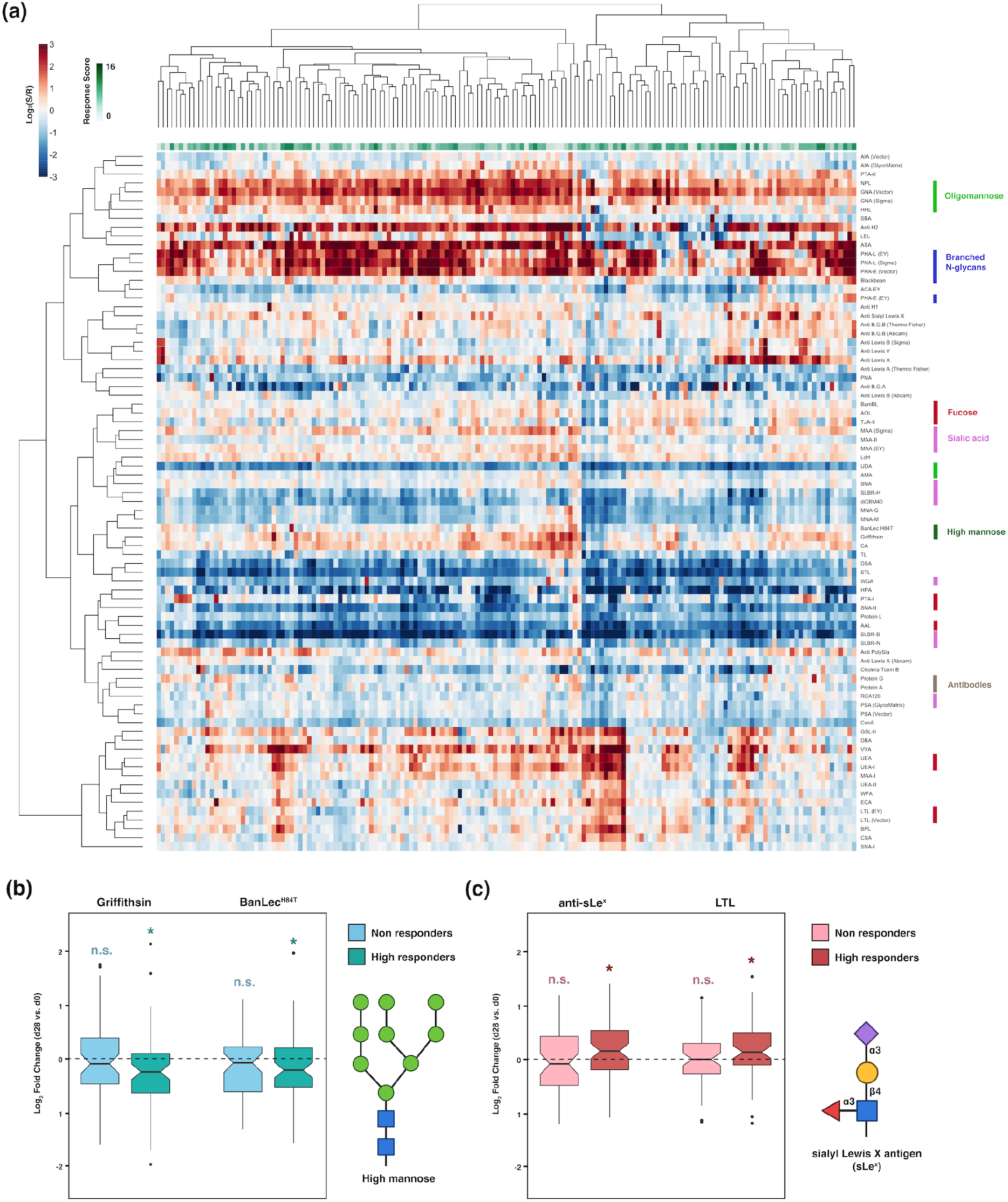
**(a)** Heatmap of lectin microarray data for post-vaccination (day 28) serum samples. Columns represent the participants and rows represent the probes. Shown is the normalized log_2_ ratios (Sample signal (*S*)/Reference signal (*R*)). Total response scores are annotated with a green-white sliding scale bar on the top of the heatmap. Rough specificities of select lectins are annotated on the right of the heatmap. **(b, c)** Boxplots of Log_2_ Fold-Change for paired d28 and d0 samples in non- and high-responders. **(b)**. High-mannose binding lectins (Griffithisin and BanLec^H84T^). **(c)** sialyl Lewis X binding probes (Anti-sLe^x^ and LTL). Paired Mann–Whitney U test was used to determine *p-*values. n.s.: not statistically significant (no difference is observed d28/d0); (*) *p* < 0.05. Glycans are shown in SNFG notation at the side of the boxplots. Symbols are defined as follows: galactose (yellow ●), N-acetylglucosamine (blue □), mannose (green), sialic acid (purple ♦), fucose (red ▴).

sLe^x^, Le^x^, and high-mannose are found on a wide range of serum glycoproteins, such as immunoglobulins and complement proteins.^38,39^ Importantly, these glycans all have clear roles in the clearance of serum/plasma glycoproteins. Le^x^-bearing glycoproteins have relatively short half-lives in serum/plasma, compared to sLe^x^-bearing glycoproteins.^40–42^ Serum proteins containing high-mannose glycans are known to be cleared from circulation at faster rates via macrophage mannose receptors.^43^ Thus, our findings, together with established knowledge of protein clearance, indicate that in response to vaccination, high responders may modulate glycosylation to alter the circulation time of serum glycoproteins.

### Glycoproteomic Identification of Serum Glycoproteins Marked by Le^a^

Our d0 data indicated that levels of Le^a^ were associated with antibody response pre-vaccination. To gain more insight into this association, we performed glycoproteomic analysis using the anti-Le^a^ antibody and BambL. In brief, we pooled the serum of all participants and performed pulldowns with either BambL or anti-Le^a^ antibody. We then analyzed the isolated proteins by mass spectrometry (**Figure 4a**). After removal of all non-glycosylated proteins and those that bound to control beads, we identified 79 glycoproteins in the BambL pulldown sample and 30 for the anti-Le^a^ pulldown. Glycoproteins enriched by the two probes are listed in **Supporting Information, Table S2 and Table S3**. Major glycoproteins enriched by BambL included immunoglobulins, complement proteins, cell adhesion molecules, protease inhibitors, and proteins in the blood coagulation pathways. As expected, the spectrum of proteins enriched by anti-Le^a^ is narrower since BambL has a broader specificity than anti-Le^a^ (**Figure 4b**). Gene ontology enrichment analysis showed enrichment for complement activation and humoral immunity in both samples. Among the pathways with more than 10-fold enrichment, the complement-related pathways had the highest number of protein hits (**Figure 4c** and **Table 1**).

**Table 1.** Complement-related Serum Glycoproteins Enriched by BambL and/or Anti-Le^a^

| Swiss-Prot accession number \| entry name | protein name | BambL | Anti-Le<sup>a</sup> |
| --- | --- | --- | --- |
| P04003 \| C4BPA_HUMAN | C4b-binding protein alpha chain | + | + |
| Q9NZP8 \| C1RL_HUMAN | Complement C1r subcomponent-like protein |  | + |
| P09871 \| C1S_HUMAN | Complement C1s subcomponent | + |  |
| P01024 \| CO3_HUMAN | Complement C3 | + |  |
| P07357 \| CO8A_HUMAN | Complement component C8 alpha chain |  | + |
| P02748 \| CO9_HUMAN | Complement component C9 |  | + |
| P00751 \| CFAB_HUMAN | Complement factor B |  | + |
| P08603 \| CFAH_HUMAN | Complement factor H | + |  |
| P05156 \| CFAI_HUMAN | Complement factor I | + |  |
| Q15485 \| FCN2_HUMAN | Ficolin-2 |  | + |
| P48740 \| MASP1_HUMAN | Mannan-binding lectin serine protease 1 | + |  |
| O00187 \| MASP2_HUMAN | Mannan-binding lectin serine protease 2 | + |  |

**Figure 4.**
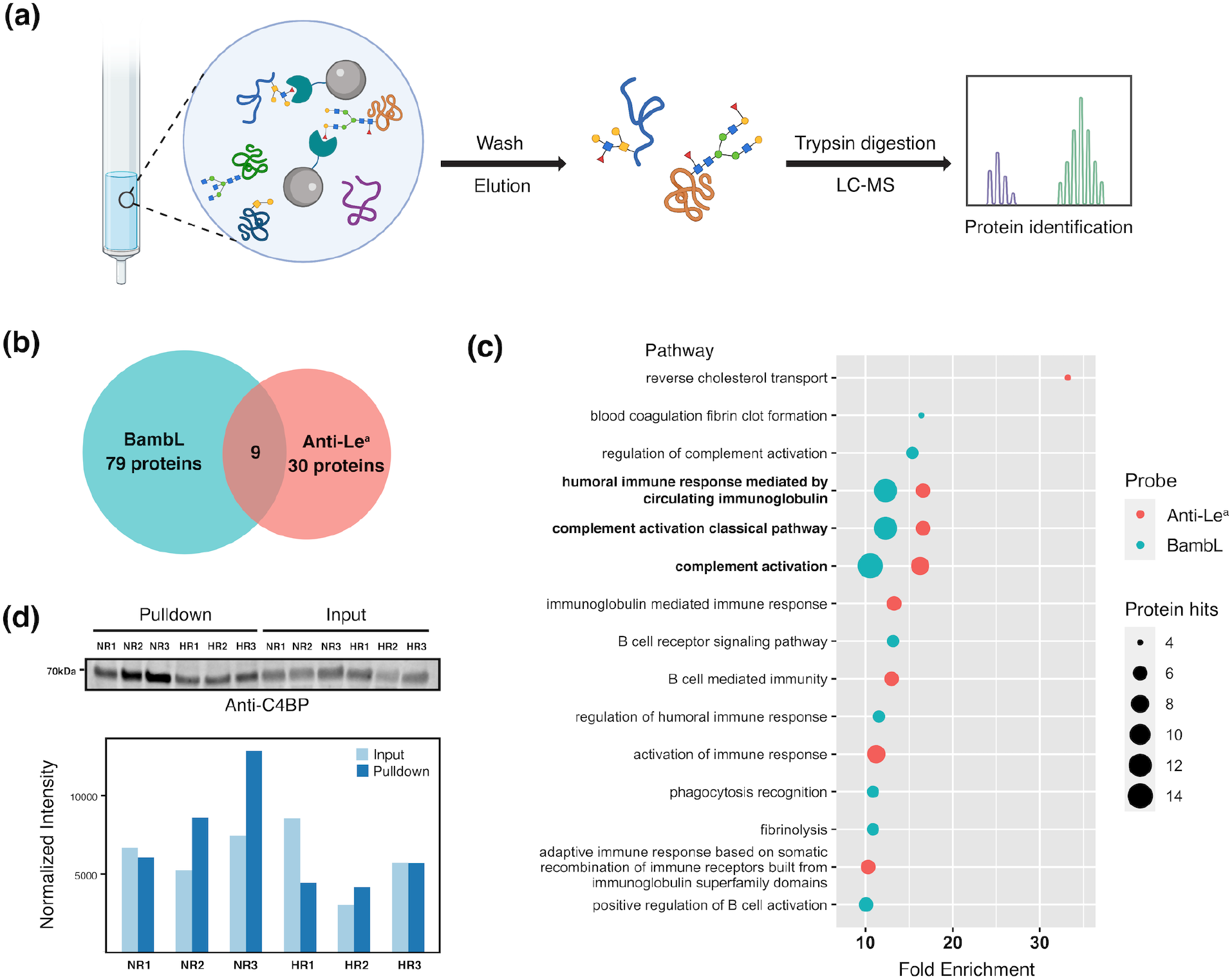
**(a)** Scheme of the experimental approach of glycoproteomic analysis. **(b)** Number of glycoproteins identified in BambL/anti-Le^a^ pulldown experiments. **(c)** Gene ontology pathway enrichment analysis for glycoproteins enriched with BambL/anti-Le^a^. The false discovery rates (FDRs) of the enriched pathways shown are all < 0.05. **(d)** Differential C4BP glycosylation. Western blot analysis for C4BP of BamBL pulldown samples and corresponding input for three high responders (HR1, HR2 and HR3) and three non-responders (NR1, NR2, and NR3) is shown. Signal intensities of the bands (normalized to total protein stain) are depicted in the bar plot.

Of the glycoproteins identified in our analysis, we selected C4b-binding protein (C4BP) for validation because it was the most abundant complement protein in our analysis and was pulled down by both BambL and anti-Le^a^. C4BP has a high serum concentration (∼ 0.2 mg/ml),^44^ thus it may have a significant contribution to the differences in BambL binding observed in lectin microarrays. For our analysis, we performed BambL-pulldowns from six individual sera samples: 3 from non-responders with high BamBL binding and 3 from high responders with low BambL binding. We then performed Western blot analysis with an anti-C4BP antibody (**Figure 4d**). As expected, Western blot analysis showed that C4BP was pulled down by BambL in all samples. In non-responders, we observed an enrichment in C4BP pulled down by BambL that was not due to a significant change in C4BP sera levels, as seen in the input samples. Our results confirm that C4BP is differentially glycosylated in non-versus high-responders.

The role of complement in vaccine efficacy is unclear. Complement is an essential aspect of both innate and adaptive immune responses and can be triggered both by immune lectins, such as MBL2 or ficolins, and by antibodies **(Figure 5)**. Studies have shown that depletion of MBL2, which is an upstream trigger for the lectin mediated complement cascade, enhances antibody production in response to both hepatitis B and tetanus toxin in mouse.^45^ In contrast, deletion of the downstream complement component C3 was found to lower antibody response to influenza antigens.^46^ There is also evidence that C2 deficiency may be associated with weak antibody response to pneumococcal vaccines.^47^ Any role glycosylation might have in the interplay between the complement cascade and antibody response has not yet been explored. Our work suggests that the glycoform of complement proteins might play a role in mediating antibody induction in response to vaccination and as a result may predict immune response to vaccines.

**Figure 5.**
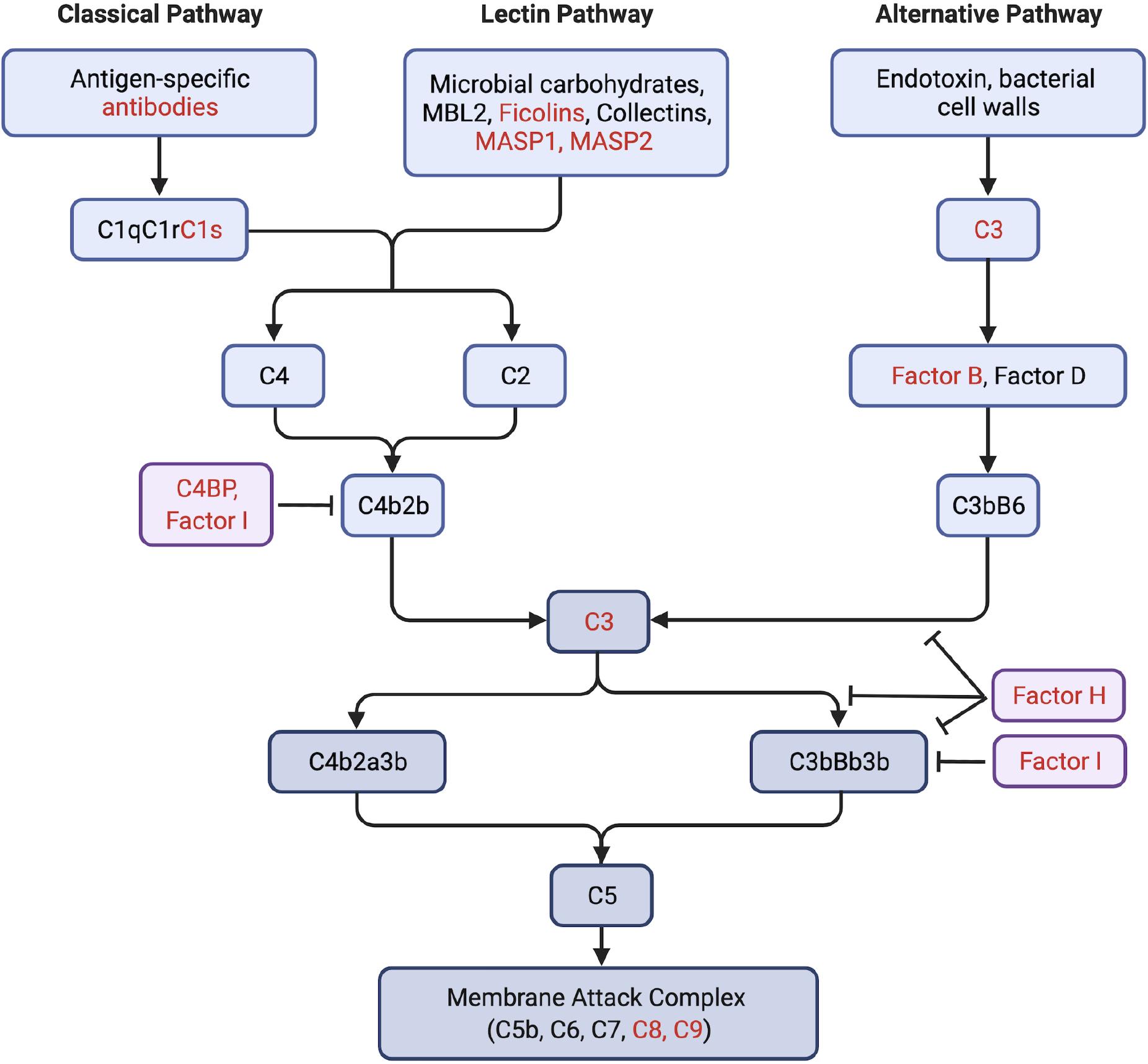
Multiple components of the complement activation pathways are glycosylated with Le^a^. For simplicity, only select components in the pathways are shown. Glycoproteins enriched by BambL or anti-Le^a^ are colored in red.

## CONCLUSION

Awareness of how vaccine effectiveness varies across populations is currently at an all-time high. The underlying reasons for such differences, however, are still opaque. Until recently, glycosylation, which is a critical modulator of immunity, has been missing from the picture. In this study, we examine the glycosylation of sera glycoproteins pre- and post-vaccination with the FLUZONE™ influenza vaccine. Our data identified high baseline levels of Le^a^ on sera glycoproteins as a potential indicator of unresponsiveness to vaccination. Glycoproteomics showed that Le^a^ was enriched in complement components such as C4BP. The role of complement in vaccine effectiveness is currently unclear. Our data argues that this may partly be due to the importance of glycoforms, which have not previously been considered. Further studies to explore how glycans such as Le^a^ influence the functions of serum glycoproteins are warranted, especially in regard to components of the complement system and the predictive ability of glycoforms to identify non-responders to specific vaccines.

## Data Availability

All data produces are either contained in the manuscript or are available online at Synapse.org (doi: 10.7303/syn26956958)

https://www.synapse.org/#!Synapse:syn26956958/files/

## ASSOCIATED CONTENT

### Supporting Information

Volcano plots comparing pre-vaccination lectin microarray data for high responders and non-responders to each strain of the influenza vaccine.

Boxplot analysis of the correlation coefficient between pre- and post-vaccination lectin microarray data.

Volcano plots comparing the paired pre- and post-vaccination lectin microarray data of all participants, high-responder only, and non-responder only. (PDF)

## AUTHOR INFORMATION

### Notes

The authors declare no competing financial interest.

## ACKNOWLEDGMENT

Some graphical contents were created with biorender.com. This project has been funded by the National Institute of Allergy and Infectious Diseases, a component of the NIH, Department of Health and Human Services, under contract 75N93019C00052.

## TOC Graphic

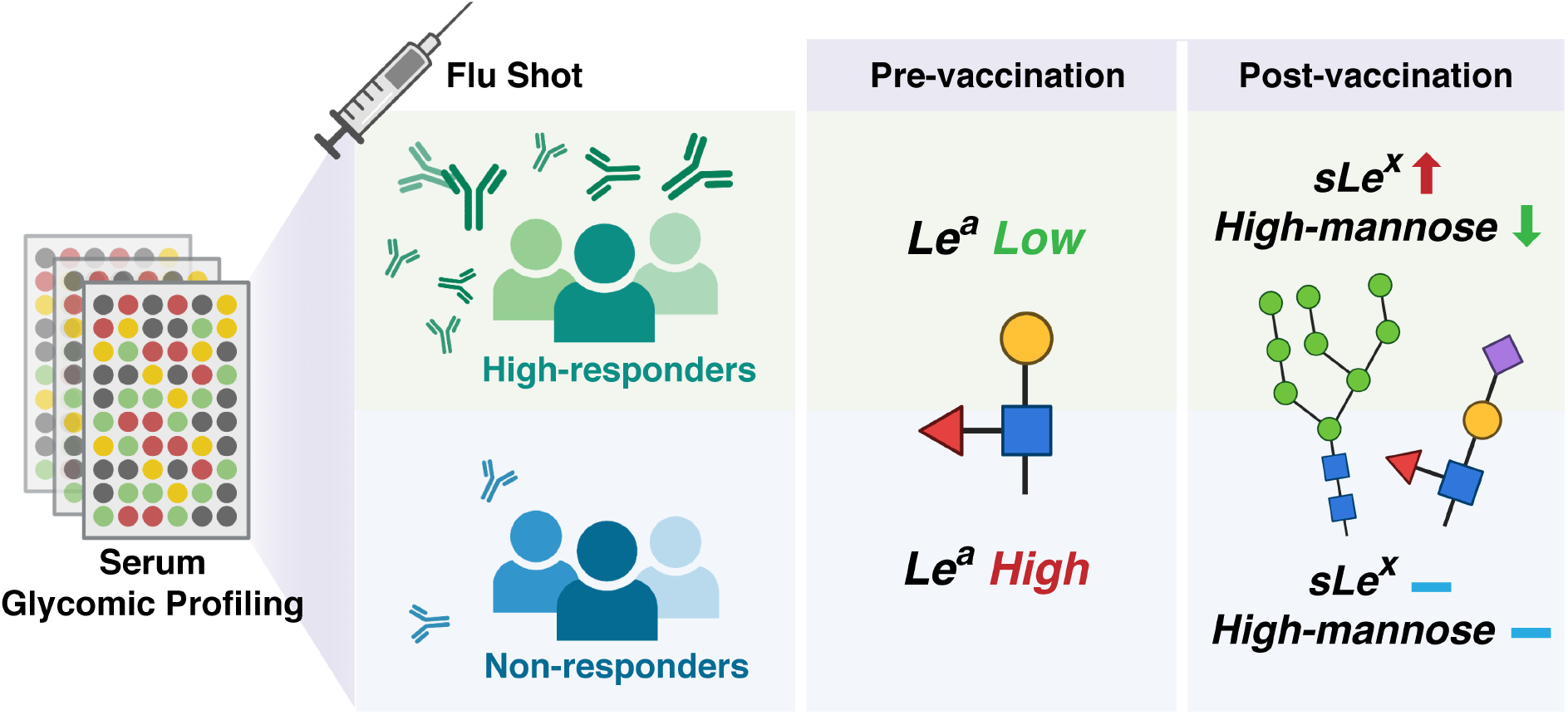

