## Supplemental Information for "Glycomic Analysis Identifies Pre-Vaccination Markers of Response to Influenza Vaccine, Implicating the Complement Pathway"

### Supporting Information

### Pre- and Post-vaccination Glycomic Profiling Reveals Association of Serum Protein Glycosylation with Antibody Responses to Influenza Vaccines

*Rui Qin[†], Guanmin Meng[†], Smruti Pushalkar[‡], Michael A. Carlock[§], Ted M. Ross[§], Christine Vogel[‡], Lara K. Mahal[†]**

† Department of Chemistry, University of Alberta, Edmonton, AB T6G 2G2, CANADA;

‡ Center for Genomics and Systems Biology, Department of Biology, New York University, New York, NY 10003, USA;

§ Center for Vaccines and Immunology, University of Georgia, Athens, GA 30602, USA

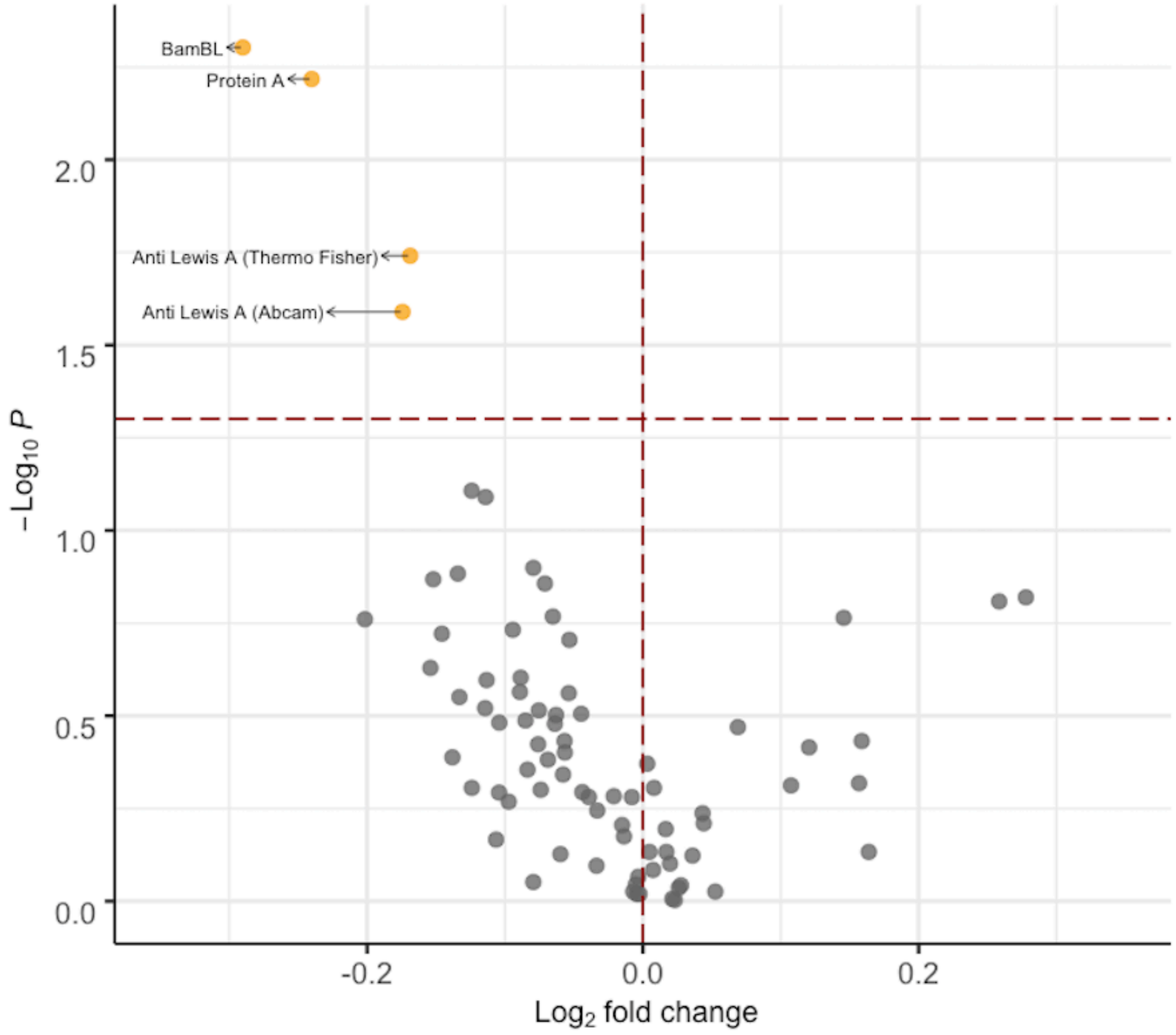


**Figure S1.** Volcano plot comparing lectin microarray data for high responders (N = 67) and non-responders (N = 65) pre-vaccination to the A/H1N1 (A/Brisbane/02/2018) strain. A positive fold change denotes higher binding of the probe in high responders. Mann–Whitney U test was used to determine p-values. Probes with $p < 0.05$ are colored in yellow.

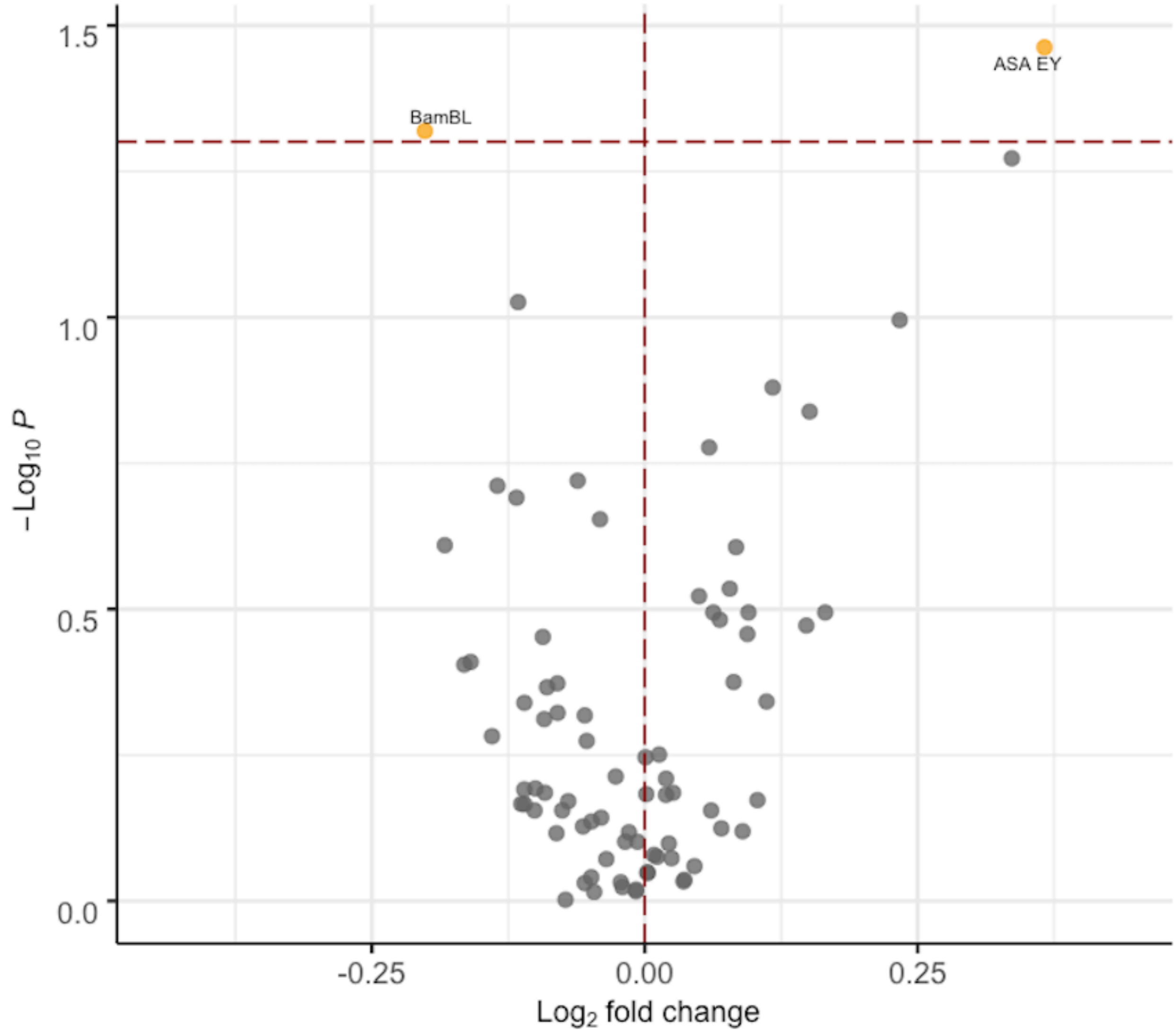


**Figure S2.** Volcano plot comparing lectin microarray data for high responders (N = 86) and non-responders (N = 64) pre-vaccination to the A/H3N2 (A/Kansas/14/2017) strain. A positive fold change denotes higher binding of the probe in high responders. Mann–Whitney U test was used to determine p-values. Probes with $p < 0.05$ are colored in yellow.

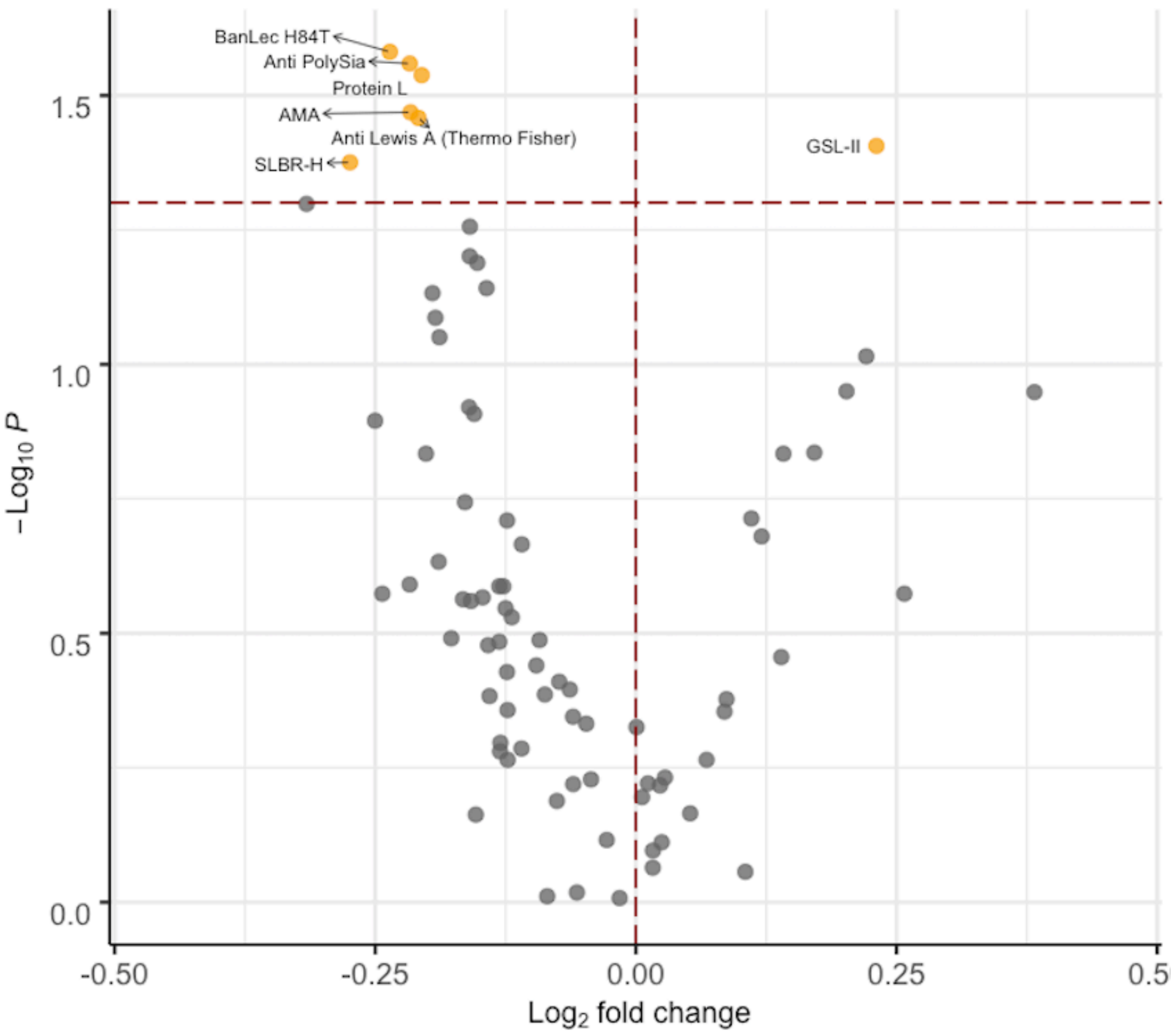


**Figure S3.** Volcano plot comparing lectin microarray data for high responders (N = 65) and non-responders (N = 62) pre-vaccination to the B/Yamagata (B/Phuket/3073/2013) strain. A positive fold change denotes higher binding of the probe in high responders. Mann–Whitney U test was used to determine p-values. Probes with $p < 0.05$ are colored in yellow.

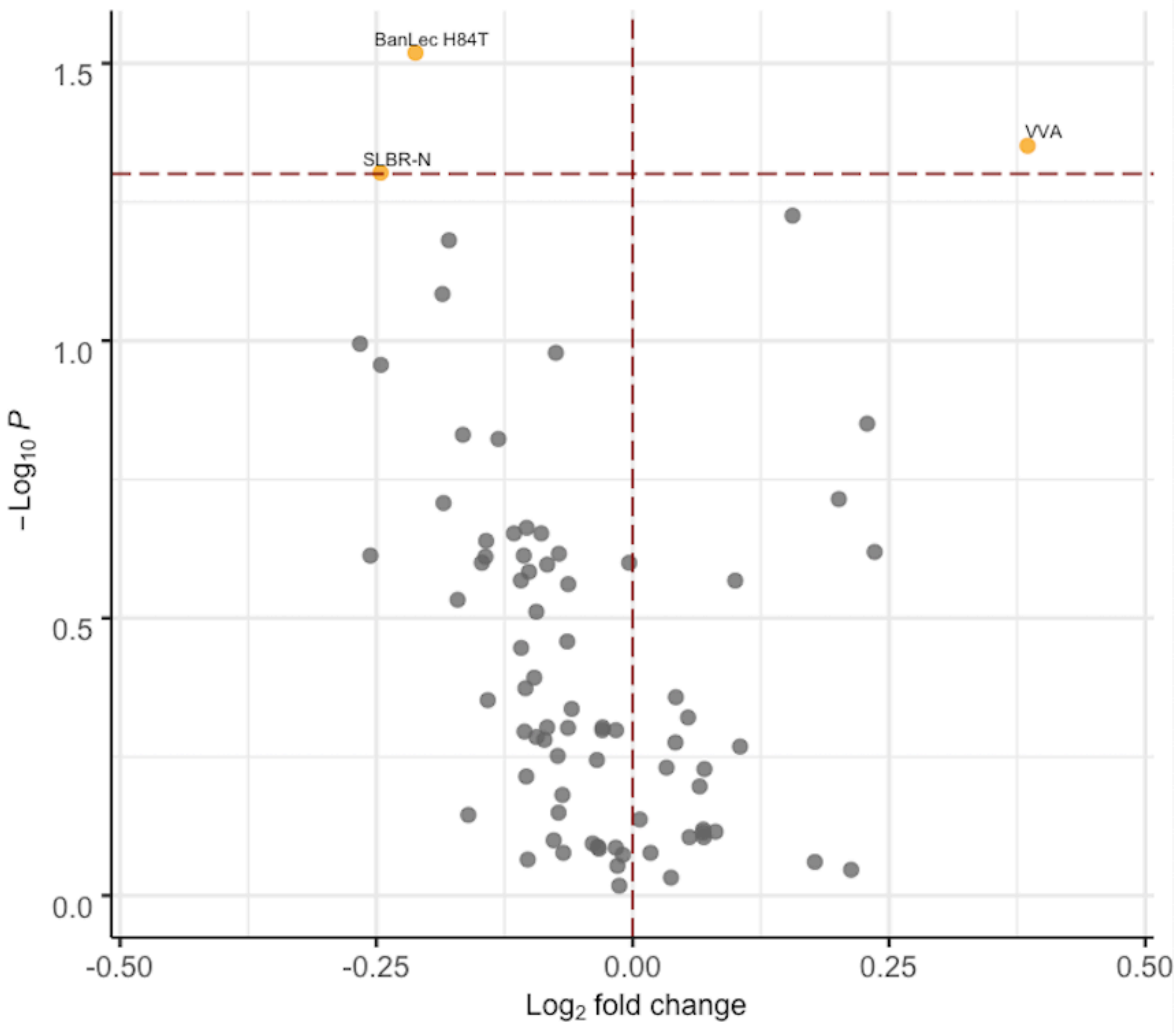


**Figure S4.** Volcano plot comparing lectin microarray data for high responders (N = 68) and non-responders (N = 65) pre-vaccination to the B/Victoria (B/Colorado/6/2017-like) strain. A positive fold change denotes higher binding of the probe in high responders. Mann–Whitney U test was used to determine p-values. Probes with $p < 0.05$ are colored in yellow.

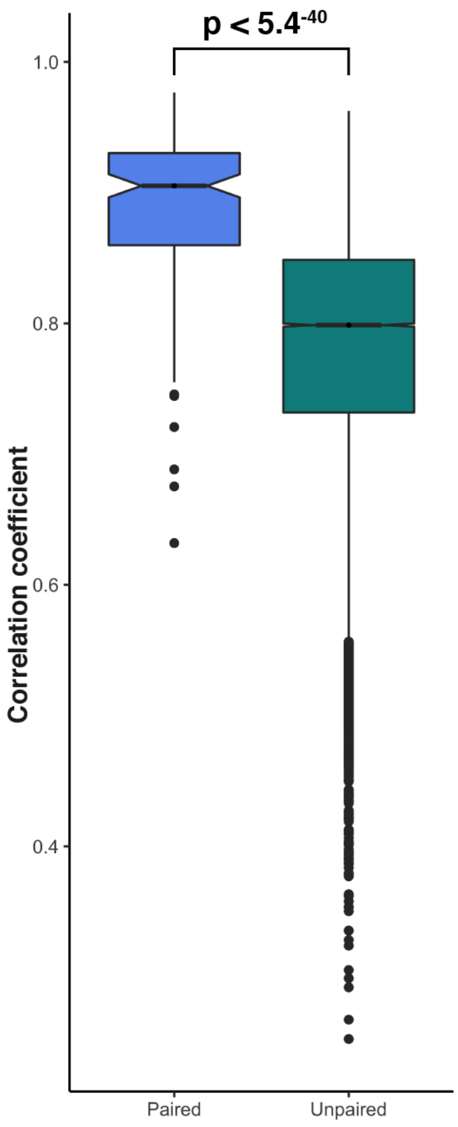


**Figure S5.** Boxplot analysis of the correlation coefficient between pre- and post-vaccination lectin microarray data. Paired: correlation coefficients of the pre- and post-vaccination glycomes of the same individuals; Unpaired: correlation coefficients of the pre- and post-vaccination glycomes of different individuals. Student's t-test was performed on the Fisher transformed-correlation coefficients.

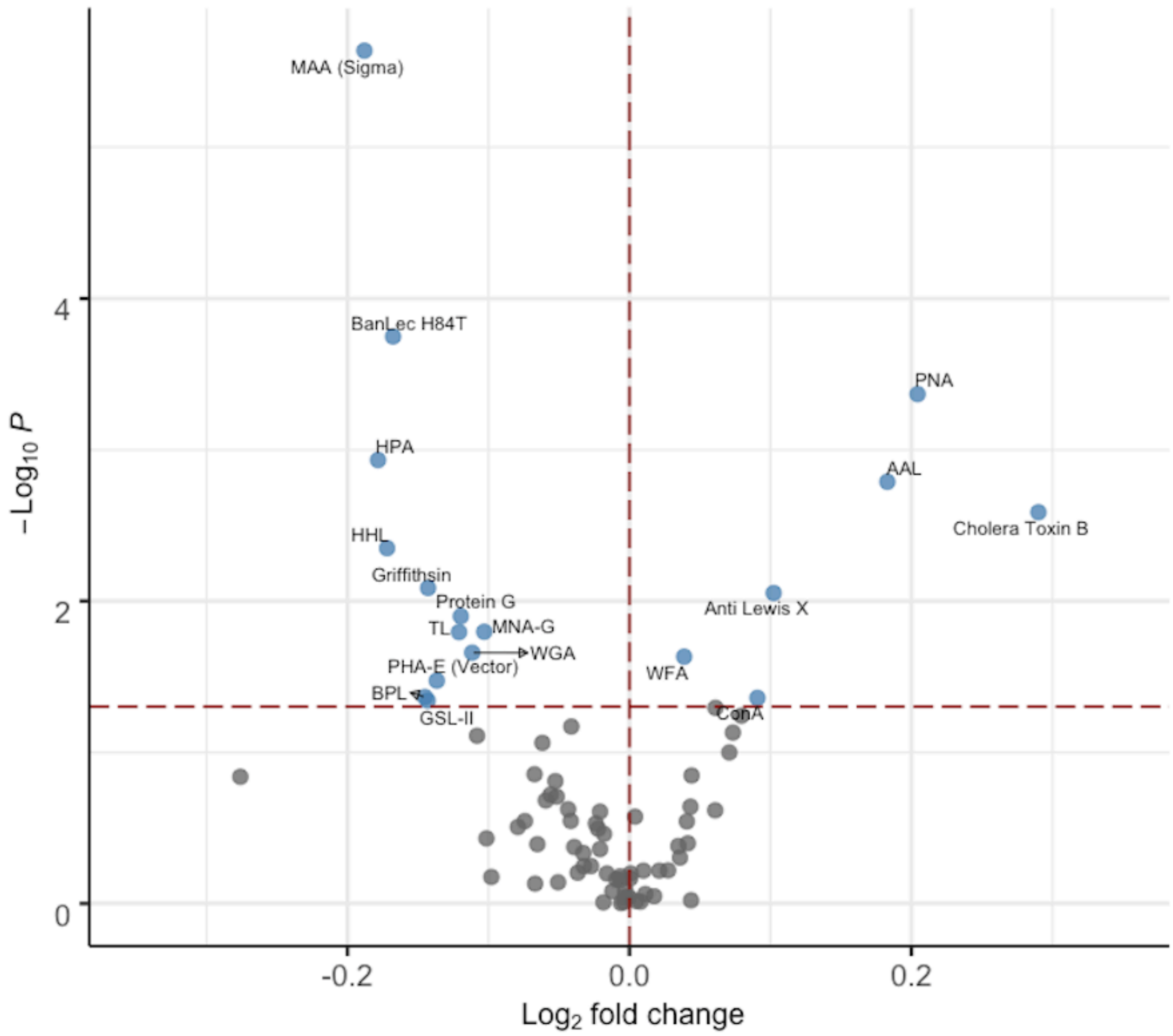


**Figure S6**. Volcano plots comparing the paired pre- and post-vaccination lectin microarray data of all participants. A positive fold change denotes higher binding of the probe to post-vaccination sera (upregulation). Paired Mann–Whitney U test was used to determine p-values. Probes with p < 0.05 are colored in blue.

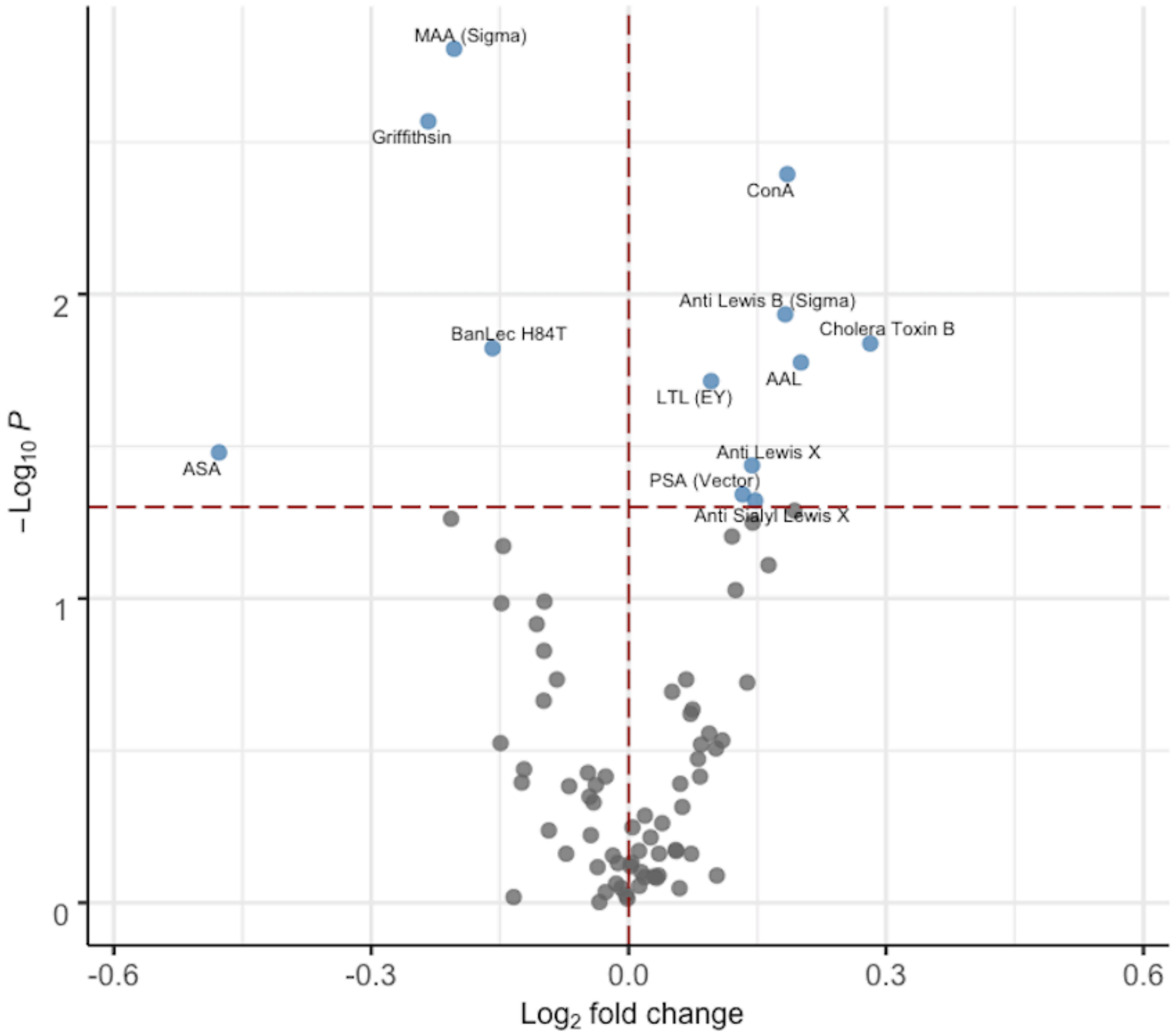


**Figure S7**. Volcano plots comparing the paired pre- and post-vaccination lectin microarray data of high responders. A positive fold change denotes higher binding of the probe to post-vaccination sera (upregulation). Paired Mann–Whitney U test was used to determine p-values. Probes with $p < 0.05$ are colored in blue.

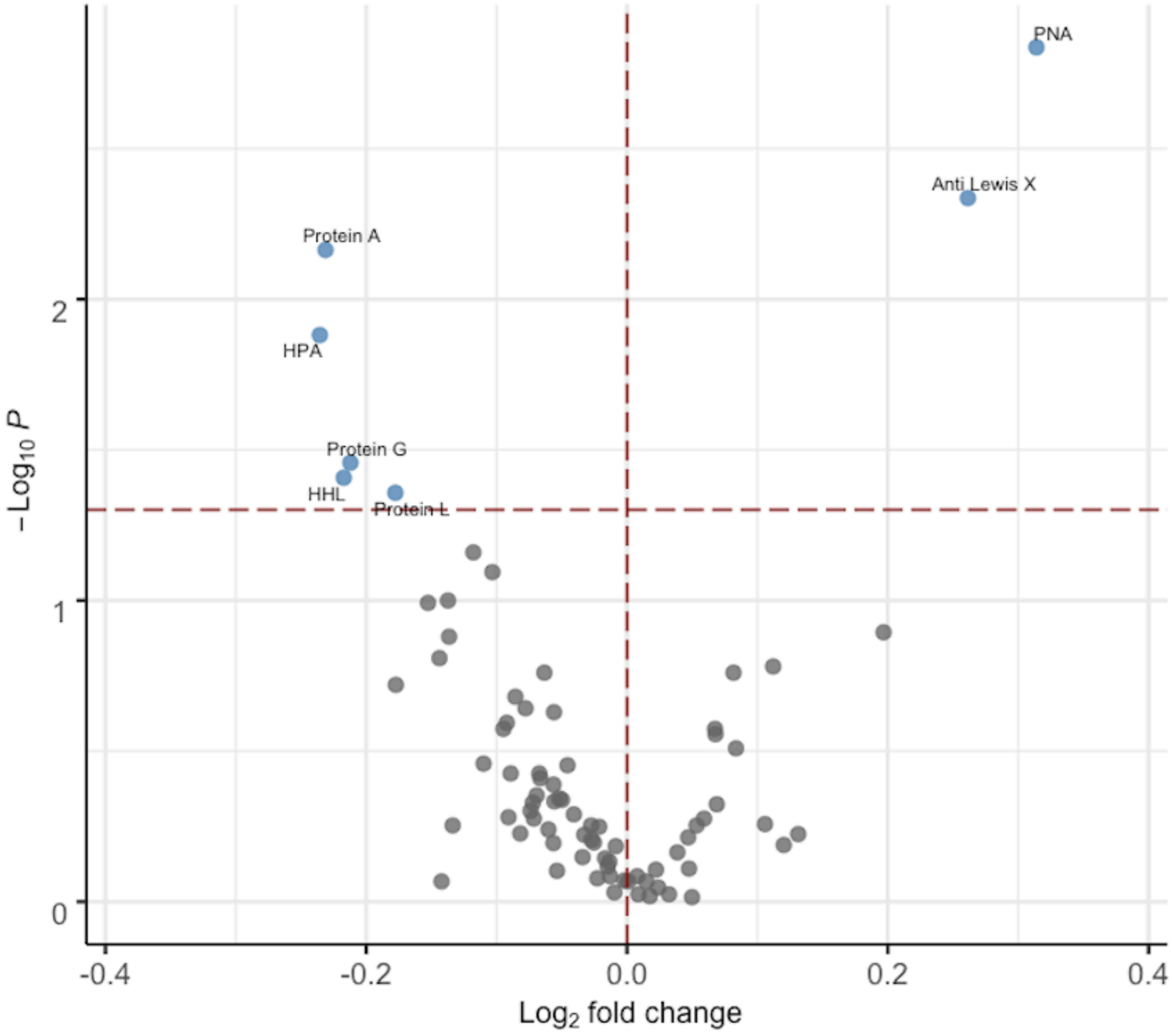


**Figure S8**. Volcano plots comparing the paired pre- and post-vaccination lectin microarray data of non-responders. A positive fold change denotes higher binding of the probe to post-vaccination sera (upregulation). Paired Mann–Whitney U test was used to determine p-values. Probes with $p < 0.05$ are colored in blue.

**Table S1.** List of Probes Printed on Lectin Microarrays

| Probe | Species of origin | Source | Printing concentration (mg/ml) | Inhibiting Sugar* |
|---|---|---|---|---|
| ConA | *Concanavalin A* | Millipore Sigma | 2 | Man |
| GNA | *Galanthus nivalis* | Millipore Sigma | 2 | Man |
| AIA | *Artocarpus integrifolia* | GlycoMatrix/Bio-world | 2 | Gal |
| PHA-E | *Phaseolus vulgaris* | Vector Labs | 2 | GlcNAc |
| AIA | *Artocarpus integrifolia* | Vector Labs | 2 | Gal |
| LcH | *Lens culinaris* | Vector Labs | 2 | Man |
| SNA | *Sambucus nigra* | Vector Labs | 2 | Lac |
| WGA | *Triticum vulgare* | Vector Labs | 2 | GlcNAc |
| PNA | *Arachis hypogaea* | Vector Labs | 2 | Gal |
| AAL | *Aleuria aurantia* | Vector Labs | 1.5 | Fuc |
| PSA | *Pisum sativum* | Vector Labs | 2 | Man |
| PSA | *Pisum sativum* | GlycoMatrix/Bio-world | 2 | Man |
| UEA-I | *Ulex europaeus* | Vector Labs | 2 | Fuc |
| LEL | *Lycopersicon esculentum* | Vector Labs | 2 | GlcNAc |
| MPL | *Machura pomifera* | Vector Labs | 2 | Gal |
| BPL | *Bauhinia purpurea* | Vector Labs | 2 | Gal |
| MAA-I | *Maachia amurensis* | Vector Labs | 2 | Lac |
| STL | *Solanum tuberosum* | Vector Labs | 2 | GlcNAc |
| WFA | *Wisteria floribunda* | Vector Labs | 2 | Gal |
| DBA | *Dolichos Biflorus* | Vector Labs | 2 | Gal |
| SBA | *Glycine max* | Vector Labs | 2 | Gal |
| HHL | *Hippeastrum hybrid* | Vector Labs | 2 | Man |
| DSA | *Datura stramonium* | Vector Labs | 2 | Lac |

| | | | | |
|---|---|---|---|---|
| ECA | *Erythrina cristagalli* | Vector Labs | 2 | GlcNAc |
| GSL-II | *Griffonia simplicifolia* | Vector Labs | 2 | GlcNAc |
| LTL | *Lotus tetragonolobus* | Vector Labs | 2 | Fuc |
| GNA | *Galanthus nivalis* | Vector Labs | 2 | Man |
| NPL | *Narcissus pseudonarcissus* | Vector Labs | 2 | Man |
| MAA-II | *Maachia amurensis* | Vector Labs | 1 | Lac |
| VVA | *Vicia villosa* | Vector Labs | 2 | Gal |
| UEA | *Ulex europaeus* | Millipore Sigma | 2 | Fuc |
| PHA-L | *Phaseolus vulgaris* | Millipore Sigma | 2 | GlcNAc |
| MAA | *Maachia amurensis* | Millipore Sigma | 2 | Lac |
| SNA | *Sambucus nigra* | Millipore Sigma | 2 | Lac |
| MNA-M | *Morniga M* | EY Labs | 2 | Man |
| SNA-I | *Sambucus nigra* | EY Labs | 2 | Lac |
| SNA-II | *Sambucus nigra* | EY Labs | 2 | Gal |
| PHA-E | *Phaseolus vulgaris* | EY Labs | 2 | GlcNAc |
| CA | *Colchicum autumnale* | EY Labs | 2 | GlcNAc |
| MNA-G | *Morniga G* | EY Labs | 2 | Gal |
| UEA-II | *Ulex europaeus* | EY Labs | 2 | GlcNAc |
| CSA | *Pure cytisus* | EY Labs | 2 | Gal |
| UDA | *Urtica dioica* | EY Labs | 2 | GlcNAc |
| LcH | *Lens culinaris* | EY Labs | 2 | Man |
| TL | *Tulipa sp.* | EY Labs | 2 | GlcNAc |
| ACA | *Amaranthus caudatus* | EY Labs | 2 | GlcNAc |
| Blackbean | *Phaseolus vulgaris* | EY Labs | 2 | Lac |
| PHA-L | *Phaseolus vulgaris* | EY Labs | 2 | GlcNAc |
| UEA-I | *Ulex europaeus* | EY Labs | 2 | Fuc |
| LTL | *Lotus tetragonolobus* | EY Labs | 2 | Fuc |

| | | | | |
|---|---|---|---|---|
| MAA | *Maachia amurensis* | EY Labs | 2 | Lac |
| PTA GalNAc | *Psophocarpus tetragonolobus* | EY Labs | 2 | Gal |
| AMA | *Arum maculatum* | EY Labs | 2 | Man |
| PTA Gal | *Psophocarpus tetragonolobus* | EY Labs | 2 | Gal |
| ASA | *Allium sativum* | EY Labs | 2 | Man |
| HPA | *Helix pomatia* | Millipore Sigma | 2 | Gal |
| TJA-II | *Trichosanthes jopanica* | Aniara | 2 | Lac |
| diCBM40 | *Clostridium perfringens* | expressed in-house | 1.5 | Lac |
| AOL | *Aspergillus oryzae* | TCI America | 2 | Fuc |
| Griffithsin | *Griffithsia* | expressed in-house | 1.7 | Man |
| SLBR-N | *Streptococcus gordonii* | expressed in-house | 2 | Lac |
| SLBR-H | *Streptococcus gordonii* | expressed in-house | 2 | Lac |
| SLBR-B | *Streptococcus gordonii* | expressed in-house | 2.3 | Lac |
| BamBL | *Burkholderia cepacia* | expressed in-house | 1.5 | Fuc |
| BanLec H84T | *Musa paradisiaca* | expressed in-house | 1 | Man |
| Protein A | | Thermo Fisher | 0.5 | / |
| Protein G | | Thermo Fisher | 1 | / |
| Protein L | | Thermo Fisher | 0.5 | / |
| Anti Lewis A | | Abcam | as received | / |
| Anti Lewis X | | Millipore Sigma | as received | / |
| Anti Lewis Y | | Abcam | as received | / |
| Anti Lewis B | | Millipore Sigma | as received | / |
| Anti Sialyl Lewis X | | GeneTex | as received | / |
| Anti H1 | | Thermo Fisher / Invitrogen | as received | / |
| Anti H2 | | Santa Cruz Biotechnology | as received | / |

| | | | | |
|---|---|---|---|---|
| Anti PolySia | | Absolute Antibody | as received | / |
| Anti Lewis A | | Thermo Fisher / Invitrogen | as received | / |
| Anti Lewis B | | Abcam | as received | / |
| Anti B.G.A | | Abcam | as received | / |
| Anti B.G.A | | Thermo Fisher / Invitrogen | as received | / |
| Anti B.G.B | | Abcam | as received | / |
| Anti B.G.B | | Thermo Fisher / Invitrogen | as received | / |
| Cholera Toxin B | *Vibrio cholerae* | Millipore Sigma | 1 | Lac |
| RCA120 | *Ricinus communis* | Vector Labs | 2 | Gal |
| Ricin B | *Ricinus communis* | Vector Labs | 1 | Lac |

*Inhibiting sugars: Man, mannose; Gal, galactose; Fuc, fucose; GlcNAc, N-acetylglucosamine; Lac, lactose.

**Table S2**. Serum Glycoproteins Enriched by BambL

| Swiss-Prot accession number | Swiss-Prot entry name | Protein name |
|---|---|---|
| P19823 | ITIH2_HUMAN | Inter-alpha-trypsin inhibitor heavy chain H2 |
| P09871 | C1S_HUMAN | Complement C1s subcomponent |
| P04003 | C4BPA_HUMAN | C4b-binding protein alpha chain |
| P01023 | A2MG_HUMAN | Alpha-2-macroglobulin |
| O95445 | APOM_HUMAN | Apolipoprotein M |
| Q06033 | ITIH3_HUMAN | Inter-alpha-trypsin inhibitor heavy chain H3 |
| P35542 | SAA4_HUMAN | Serum amyloid A-4 protein |
| Q96PD5 | PGLYRP2_HUMAN | N-acetylmuramoyl-L-alanine amidase |
| P01876 | IGHA1_HUMAN | Immunoglobulin heavy constant alpha 1 |
| P01857 | IGHG1_HUMAN | Immunoglobulin heavy constant gamma 1 |
| P23142 | FBLN1_HUMAN | Fibulin-1 |
| P02765 | FETUA_HUMAN | Alpha-2-HS-glycoprotein |
| P01859 | IGHG2_HUMAN | Immunoglobulin heavy constant gamma 2 |
| P05156 | CFAI_HUMAN | Complement factor I |
| P10909 | CLUS_HUMAN | Clusterin |
| P14151 | LYAM1_HUMAN | L-selectin |
| P01024 | CO3_HUMAN | Complement C3 |
| P20742 | PZP_HUMAN | Pregnancy zone protein |
| P01877 | IGHA2_HUMAN | Immunoglobulin heavy constant alpha 2 |
| Q7Z5P9 | MUC19_HUMAN | Mucin-19 |
| P08603 | CFAH_HUMAN | Complement factor H |
| P02749 | APOH_HUMAN | Beta-2-glycoprotein 1 |
| P02743 | SAMP_HUMAN | Serum amyloid P-component |
| P02671 | FIBA_HUMAN | Fibrinogen alpha chain |

| | | |
|---|---|---|
| P13598 | ICAM2_HUMAN | Intercellular adhesion molecule 2 |
| P51884 | LUM_HUMAN | Lumican |
| P00747 | PLMN_HUMAN | Plasminogen |
| P05546 | HEP2_HUMAN | Heparin cofactor 2 |
| Q96IY4 | CBPB2_HUMAN | Carboxypeptidase B2 |
| O75882 | ATRN_HUMAN | Attractin |
| P08519 | APOA_HUMAN | Apolipoprotein(a) |
| P05090 | APOD_HUMAN | Apolipoprotein D |
| P05452 | TETN_HUMAN | Tetranectin |
| Q6UXB8 | PI16_HUMAN | Peptidase inhibitor 16 |
| Q9UK55 | ZPI_HUMAN | Protein Z-dependent protease inhibitor |
| Q14520 | HABP2_HUMAN | Hyaluronan-binding protein 2 |
| P43121 | MUC18_HUMAN | Cell surface glycoprotein MUC18 |
| Q12913 | PTPRJ_HUMAN | Receptor-type tyrosine-protein phosphatase eta |
| P08637 | FCG3A_HUMAN | Low affinity immunoglobulin gamma Fc region receptor III-A |
| P22105 | TENX_HUMAN | Tenascin-X |
| P33151 | CADH5_HUMAN | Cadherin-5 |
| P07333 | CSF1R_HUMAN | Macrophage colony-stimulating factor 1 receptor |
| P26927 | HGFL_HUMAN | Hepatocyte growth factor-like protein |
| P15151 | PVR_HUMAN | Poliovirus receptor |
| P13473 | LAMP2_HUMAN | Lysosome-associated membrane glycoprotein 2 |
| P15814 | IGLL1_HUMAN | Immunoglobulin lambda-like polypeptide 1 |
| P01833 | PIGR_HUMAN | Polymeric immunoglobulin receptor |
| Q9HDC9 | APMAP_HUMAN | Adipocyte plasma membrane-associated protein |
| P55290 | CAD13_HUMAN | Cadherin-13 |
| P01033 | TIMP1_HUMAN | Metalloproteinase inhibitor 1 |
| P48740 | MASP1_HUMAN | Mannan-binding lectin serine protease 1 |
| Q6YHK3 | CD109_HUMAN | CD109 antigen |

| | | |
|---|---|---|
| P41222 | PTGDS_HUMAN | Prostaglandin-H2 D-isomerase |
| Q9UNN8 | EPCR_HUMAN | Endothelial protein C receptor |
| O00533 | NCHL1_HUMAN | Neural cell adhesion molecule L1-like protein |
| P12821 | ACE_HUMAN | Angiotensin-converting enzyme |
| P10153 | RNAS2_HUMAN | Non-secretory ribonuclease |
| P13591 | NCAM1_HUMAN | Neural cell adhesion molecule 1 |
| P35916 | VGFR3_HUMAN | Vascular endothelial growth factor receptor 3 |
| O00187 | MASP2_HUMAN | Mannan-binding lectin serine protease 2 |
| P12830 | CDH1_HUMAN | Cadherin-1 |
| P10721 | KIT_HUMAN | Mast/stem cell growth factor receptor Kit |
| P62937 | PPIA_HUMAN | Peptidyl-prolyl cis-trans isomerase A |
| P16070 | CD44_HUMAN | CD44 antigen |
| P16109 | LYAM3_HUMAN | P-selectin |
| P12259 | FAS_HUMAN | Coagulation factor V |
| P05164 | PERM_HUMAN | Myeloperoxidase |
| P17813 | EGLN_HUMAN | Endoglin |
| Q6UX71 | PLXDC2_HUMAN | Plexin domain-containing protein 2 |
| P05154 | IPSP_HUMAN | Plasma serine protease inhibitor |
| P02786 | TFR1_HUMAN | Transferrin receptor protein 1 |
| P07359 | GP1BA_HUMAN | Platelet glycoprotein Ib alpha chain |
| Q15762 | CD226_HUMAN | CD226 antigen |
| Q9BY67 | CADM1_HUMAN | Cell adhesion molecule 1 |
| Q8N6C8 | LILRA3_HUMAN | Leukocyte immunoglobulin-like receptor subfamily A member 3 |
| P02675 | FIBB_HUMAN | Fibrinogen beta chain |
| Q15166 | PON3_HUMAN | Serum paraoxonase/lactonase 3 |
| P17936 | IBP3_HUMAN | Insulin-like growth factor-binding protein 3 |
| Q8NBP7 | PCSK9_HUMAN | Proprotein convertase subtilisin/kexin type 9 |

**Table S3**. Serum Glycoproteins Enriched by Anti-Le[a]

| Swiss-Prot accession number | Swiss-Prot entry name | Protein name |
|---|---|---|
| P08697 | A2AP_HUMAN | Alpha-2-antiplasmin |
| P10586 | PTPRF_HUMAN | Receptor-type tyrosine-protein phosphatase F |
| Q99784 | NOE1_HUMAN | Noelin |
| P51884 | LUM_HUMAN | Lumican |
| P08519 | APOA_HUMAN | Apolipoprotein(a) |
| P19823 | ITIH2_HUMAN | Inter-alpha-trypsin inhibitor heavy chain H2 |
| P40197 | GPV_HUMAN | Platelet glycoprotein V |
| Q92820 | GGH_HUMAN | Gamma-glutamyl hydrolase |
| P02774 | VTDB_HUMAN | Vitamin D-binding protein |
| P02751 | FINC_HUMAN | Fibronectin |
| Q15485 | FCN2_HUMAN | Ficolin-2 |
| P12259 | FA5_HUMAN | Coagulation factor V |
| Q02413 | DSG1_HUMAN | Desmoglein-1 |
| P07858 | CATB_HUMAN | Cathepsin B |
| P35222 | CTNB1_HUMAN | Catenin beta-1 |
| Q9NQ79 | CRAC1_HUMAN | Cartilage acidic protein 1 |
| P10909 | CLUS_HUMAN | Clusterin |
| P00751 | CFAB_HUMAN | Complement factor B |
| P11597 | CETP_HUMAN | Cholesteryl ester transfer protein |
| P02748 | CO9_HUMAN | Complement component C9 |
| P07357 | CO8A_HUMAN | Complement component C8 alpha chain |
| P04003 | C4BPA_HUMAN | C4b-binding protein alpha chain |
| Q9NZP8 | C1RL_HUMAN | Complement C1r subcomponent-like protein |
| P06276 | CHLE_HUMAN | Cholinesterase |

| | | |
|---|---|---|
| O95445 | APOM_HUMAN | Apolipoprotein M |
| P02749 | APOH_HUMAN | Beta-2-glycoprotein 1 |
| P02656 | APOC3_HUMAN | Apolipoprotein C-III |
| P02765 | FETUA_HUMAN | Alpha-2-HS-glycoprotein |
| P43652 | AFAM_HUMAN | Afamin |
| P0DP01 | HV108_HUMAN | Immunoglobulin heavy variable 1-8 |
